# The Nature and Origins of Loneliness amongst NHS Talking Therapies Clients— A Qualitative Study from Therapists’ Perspective

**DOI:** 10.1101/2025.11.15.25340306

**Authors:** Cheuk Hei Peony Chung, Brynmor Lloyd-Evans, Sampath Rajapakse, Theodora Stefanidou

## Abstract

**Background:** Depressive and anxiety disorders are the most common mental health conditions. In the United Kingdom (UK), NHS Talking Therapies for anxiety and depression (TTad) has provided evidence-based interventions for these conditions since 2008, although treatment outcomes are often suboptimal. Loneliness, consistently linked with poorer mental health outcomes, remains unexplored in this context. We investigated the views of NHS TTad therapists about the nature and origins of loneliness amongst their clients.

**Methods:** We conducted semi-structured qualitative interviews with 19 NHS TTad therapists, purposively sampled across job roles, experience levels, and service locations. Recruitment was via professional networks and online platforms. Interviews were conducted online, and data were analysed using thematic analysis with both inductive and deductive approaches.

**Findings:** Four key themes emerged from analysis: (1) Life-transitions, (2) Mental health conditions, (3) Barriers to connection, (4) Stigma. NHS TTad therapists described loneliness experienced amongst their clients as arising from both external factors (e.g., relocation, financial difficulties) and internal psychological processes (e.g., low self-esteem, mental health conditions). Therapists believed that loneliness was experienced even when clients had social contacts, due to barriers like cultural differences and mental health symptoms. Therapists also observed some demographic variability in the experience of loneliness amongst clients (e.g., across age, gender, mental health diagnosis). Societal factors including stigma surrounding mental health and loneliness were viewed as further contributing to clients’ loneliness experiences and help-seeking behaviours.

**Conclusions:** NHS TTad therapists perceived loneliness as a common experience amongst their clients, linked to life transitions, mental health symptoms, structural, cultural, and psychological barriers to connection. These findings highlighted the importance of recognising loneliness as a relevant factor in therapy and of developing strategies to identify and address it within, or outside of NHS TTad services. Future research should also examine the impact of social media, gender, and public stigma on loneliness. Addressing these factors at societal and policy levels is essential to reduce stigma and improve support for individuals experiencing loneliness.

## Introduction

### Background

Depressive and anxiety disorders are the most common mental disorders globally, responsible for 56.3 million and 42.5 million Disability-Adjusted Life Years (DALYs) in 2021 (1,2) . DALYs measure disease burden, including years lost to premature death and years lived with disability (3). In the UK, these disorders contribute 40% and 21% of mental health DALYs, respectively (4). With 1 in 6 English individuals affected by depression and/or anxiety, these numbers are expected to rise (5).

In response, NHS Talking Therapies for anxiety and depression (TTad) (previously known as Improving Access to Psychological Therapies - IAPT) was established in England in 2008 (6). It is a primary care psychological therapy programme that uses a stepped-care model, offering evidence-based interventions tailored to clients’ needs. Psychological Wellbeing Practitioners (PWPs) provide low-intensity, cognitive-behavioural therapy (CBT)-based self-help for mild to moderate cases, whilst high-intensity therapists offer formal CBT, interpersonal therapy, and couples counselling for more severe cases (7).

In 2024-2025, 1.81 million individuals were referred to NHS TTad, with 1.21 million accessing the service (8). Despite this, only half (50.5%) of those completing treatment recovered, and only 55.5% of individuals accessed the service completed their treatment course (8).

Previous research identified factors affecting engagement, including age (9), ethnicity (10), social deprivation (11), low motivation to change (12). Moreover, factors like high baseline symptomatology, long waiting time (13), high number of missed appointments (11), comorbidity with personality disorders (14), and psychological trauma (15) are linked with poor treatment response. Nonetheless, the role of loneliness, which may influence NHS TTad treatment outcomes has not been previously explored.

### Loneliness and its relationship with depression and anxiety

Loneliness is an aversive state arising from a discrepancy between desired and perceived interpersonal relationships (16). Loneliness can be understood as a multidimensional construct with three primary dimensions: intimate, relational, and collective loneliness (17). Intimate loneliness stems from lacking emotional support from close others, such as a spouse, with marital status being a strong predictor in middle-aged and older adults (18). Relational loneliness reflects a deficiency in quality friendships or family connections (19), while collective loneliness involves a lack of community or shared identity, often linked to participation in voluntary groups (18). All three types of loneliness were found to be associated with depressive and anxiety symptoms in young adults (20–22), whereas both intimate and relational loneliness predicted depressive symptoms in older adults (23).

In a large UK population-based cross-sectional study, loneliness was found to be associated with all mental disorders, most strongly with phobia and depression (24). Later, two meta-analyses of both cross-sectional and longitudinal studies found moderately significant effects of loneliness on depression and anxiety (18,19). Moreover, in a systematic review (27), amongst the 13 papers assessed that used depression severity as an outcome, 11 of them reflected that poorer perceived social support or greater loneliness at baseline were significant predictors of greater depressive symptom severity at follow-ups. For anxiety, studies found loneliness and low perceived social support predicted anxiety symptoms (28,29). A meta-analysis also indicated that loneliness predicted social anxiety symptoms in children and adolescents (30).

Loneliness not only predicts depression and anxiety but is also exacerbated by these conditions. A longitudinal study (29) found higher social anxiety to predict greater loneliness six months later in adults. A meta-analysis also indicated social anxiety predicted future loneliness in children and adolescents (30). Similarly, depression was found to predict loneliness (31,32). Depressed individual may have distorted views of the self and others, show heightened rejection sensitivity (33), and display social disengagement (34), contributing to loneliness. Nonetheless, some studies failed to find the predictive nature of depression on loneliness (29,35) suggesting the complex relationship between loneliness and depression that requires further investigation.

### Current qualitative evidence on loneliness

Recent qualitative research has begun to explore the nuanced experiences of loneliness among individuals with mental disorders. For example, Birken et al. (36) found that loneliness in adults with various mental disorders arose from a lack of meaningful social connections, influenced by both internal (e.g., introversion) and external factors (e.g., loss and separation).

This study also highlighted the bidirectional relationship between loneliness and mental disorders, with each exacerbating the other.

Other studies have focused on specific populations, such as those with severe mental illnesses or personality disorders. Ludwig et al. (37) found that individuals with schizophrenia experienced loneliness due to factors like financial constraints, poor relationship quality, social anxiety, and internalised stigma. Similarly, Ikhtabi et al.(38)’s meta-synthesis found that loneliness associated with personality disorders is often deepened by past rejection and stigma. Symptoms of personality disorders and loneliness also reinforced one another.

In the context of depression, Achterbergh et al. (39) conducted a meta-synthesis of studies on loneliness among young people with depression. The findings indicate that depressive symptoms, such as low motivation and energy, led to social withdrawal. Their fear of judgement, losing friendships and difficulties in expressing themselves also played a role. Paradoxically, these young people expressed a desire for connection, despite their avoidance behaviours.

### Research Gaps and The Current Study

Current literature reveals strong reciprocal links between loneliness, depression, and anxiety. However, no study has explored how loneliness is experienced by NHS TTad clients with these conditions. Given the low engagement and recovery rates among these clients (8), examining loneliness as a potential factor is crucial.

Whilst quantitative studies show robust relationships between loneliness and depression/anxiety, single-item or brief measures fail to capture the full complexity of loneliness and what may precipitate or maintain it (36). Thus, this paper adopts a qualitative approach to investigate the nature and origins of loneliness among NHS TTad clients.

Current qualitative research highlights the complexity of loneliness in individuals with mental disorders. However, there is a gap in studies focusing exclusively on those with depression and anxiety, despite these being the most common mental disorders. A meta-synthesis on loneliness in individuals with depression also only focused on young people (40), leaving adult experiences unexplored.

This study therefore aimed to explore therapists’ perspectives on the nature and origins of loneliness amongst NHS TTad clients. Therapists, through their clinical training and ongoing contact with diverse clients, are uniquely positioned to identify patterns, meanings, and perceived origins of loneliness in this population (42).

## Methods

The methods and results of this study were reported following the Consolidated Criteria for Reporting Qualitative Research (COREQ) (see Appendix A).

### Ethics

Ethical approval was obtained from the University College London Research Ethics Committee (Ref:8855.001) before the commencement of recruitment and data collection. Written or verbal consent was obtained from all participants.

### Setting

This study took place in England, across various NHS trusts. Participants ultimately recruited were based in seven NHS TTad services delivered by six London NHS Trusts. These services operate in both inner and outer London boroughs that are socially and ethnically diverse.

### Participants

Purposive sampling was employed to gain in-depth insights into therapists’ perspectives on clients’ experiences of loneliness. Eligible participants were aged 18 or older, actively providing interventions to NHS TTad clients with anxiety and depression, and fluent in English. We aimed to recruit therapists representing a range of ethnic backgrounds, job roles (e.g., CBT therapists, Psychological Wellbeing Practitioners), levels of experience in NHS TTad services, and geographical locations across London. Based on prior research, we anticipated that 9–17 interviews would be sufficient to reach theoretical saturation (41).

### Recruitment

Recruitment posts were distributed within the research team’s professional network, including the North and Central East London IAPT Service Improvement and Research Network and UCL Division of Psychiatry MSc alumni. A recruitment flyer was also posted on LinkedIn, in a UCL PWP alumni WhatsApp group, and emailed to the British Psychological Society and Talking Therapies National Networking Forums for further dissemination. Interested potential participants were asked to contact the research team.

Potential participants received an information sheet and were asked to provide electronic informed consent before the interview. One participant withdrew before the interview due to time constrains and their personal data were removed from the study records. Participants received a £20 Love2shop e-voucher as compensation.

### Data collection

A topic guide (Appendix B) was developed by researcher TS, based on existing literature, consultations with the research team, and input from a stakeholders advisory group (consisting of individuals with lived experience of loneliness and depression and anxiety). The topic guide addressed three main areas: (1) the origins and extent of loneliness experienced by clients, (2) staff responses to support lonely clients, and (3) how loneliness might impact treatment engagement. This study is part of a wider research project (The Loneliness Study), a mixed methods investigation examining the impact of loneliness on psychological treatment outcomes for adults with anxiety and depression disorders. Data used for the current study mostly came from the first part of the topic guide, with sample questions like “*Is loneliness an issue for many of your clients?”*, *“What are the biggest factors that clients experience loneliness?”, “Are certain client groups more prone to loneliness?”*.

All interviews were conducted between January 2024 and December 2024. Upon consent, participants were interviewed online via Microsoft Teams by researchers TS or PC. Participants first provided demographic details (gender, age, ethnicity), their job role, NHS TTad organisation, and the length they have worked in NHS TTad and the mental health field. Semi-structured interviews were then conducted, with occasional follow-up questions asked to clarify or further explore participants’ accounts. Interviews lasted between 35 and 63 minutes (median = 51 minutes), were all video-recorded, and transcribed verbatim by an external company. All transcripts were anonymised, with identifying information removed and stored on the secure UCL online system.

### Data analysis

Thematic analysis (42) was employed to analyse the interview data, using NVivo 14 software. Primary researcher PC analysed all 19 transcripts, developed an initial coding framework and some initial themes. A second researcher (SR), coded two interviews separately, and subsequently discussed the initial coding framework with researcher PC. Two researchers BLE and TS, who are part of the research team, also provided feedback on the coding framework and initial themes upon reading two transcripts.

### Positionality and reflexivity

The researcher’s positionality could affect data collection and analysis (43). Researcher and MSc student PC (female, Asian Chinese) has a psychology and mental health research background, with experience providing low-intensity psychological intervention in Hong Kong. Her experience providing low-intensity intervention might lead to preconceptions about loneliness experienced amongst clients, although having worked in a context different from NHS TTad. BLE, Professor of Mental Health and Social Inclusion (male, White British, with a social work background) and TS, PhD candidate and Research Fellow (female, White European, with a psychology background) have expertise in loneliness research. Their backgrounds might influence data analysis and topic guide generation, introducing deductive reasoning. SR, MSc student (Male, British South Asian) has a background in mental health and psychology, with no previous experience in loneliness research.

Given the positionalities of researchers, a reflexive approach was maintained throughout the research (43). PC received training and guidance on qualitative interviewing from TS, an experienced mixed methods researcher. Data analysis involved collaborative team discussions to enhance interpretative depth, and SR, not part of the loneliness research team, also coded and reviewed frameworks for additional reflexivity. Research diaries were kept throughout to document and address reflexivity issues.

## Results

Nineteen NHS TTad therapists were interviewed, the majority of whom were female (89.5%) and aged between 25 and 63 years. Ethnically, participants were most commonly White British (36.8%) and Asian Chinese (21.1%). The distribution of low-intensity (52.6%) and high-intensity (47.4%) therapists was balanced. Participants were drawn from a range of London NHS TTad services. On average, therapists had worked 8.24 years (SD = 8.96) in mental health services and 4.95 years (SD = 4.85) within NHS TTad services. *Table 1* provides a detailed summary of these characteristics.

**Table 1.** Demographic and Professional Characteristics of Participants.

| Characteristic | Participants (N = 19) |
| --- | --- |
| **Gender - N (%)** |  |
| Female | 17 (89.5) |
| Male | 2 (10.5) |
| **Age group - N (%)** |  |
| 25-34 | 12 (63.2) |
| 35-44 | 4 (21.1) |
| 45-54 | 1 (5.3) |
| 55-64 | 2 (10.5) |
| **Ethnicity - N (%)** |  |
| White British | 7 (36.8) |
| White Other | 3 (15.8) |
| Asian Chinese | 4 (21.1) |
| Asian Other | 2 (10.5) |
| Mixed Ethnic Group | 3 (15.8) |
| **Job role - N (%)** |  |
| *High-intensity therapists* | 9 (47.4) |
| CBT Therapist | 3 (15.8) |
| Clinical Psychologist | 2 (10.5) |
| Counselling Psychologist | 1 (5.3) |
| Trainee CBT Therapist | 1 (5.3) |
| Trainee Clinical Psychologist | 2 (10.5) |
| *Low-intensity therapists* | 10 (52.6) |
| Psychological Wellbeing Practitioner | 10 (52.6) |
| **Years worked in Mental Health Services - Mean (SD)** | 8.24 (8.96) |
| **Years worked in NHS TTad service - Mean (SD)** | 4.95 (4.85) |

Thematic analysis identified four overarching themes, (1) Life transitions, (2) Mental Health Conditions, (3) Barriers to Connection, and (4) Stigma. These four themes and their corresponding subthemes are illustrated in *Figure 1* below.

**Figure 1.**
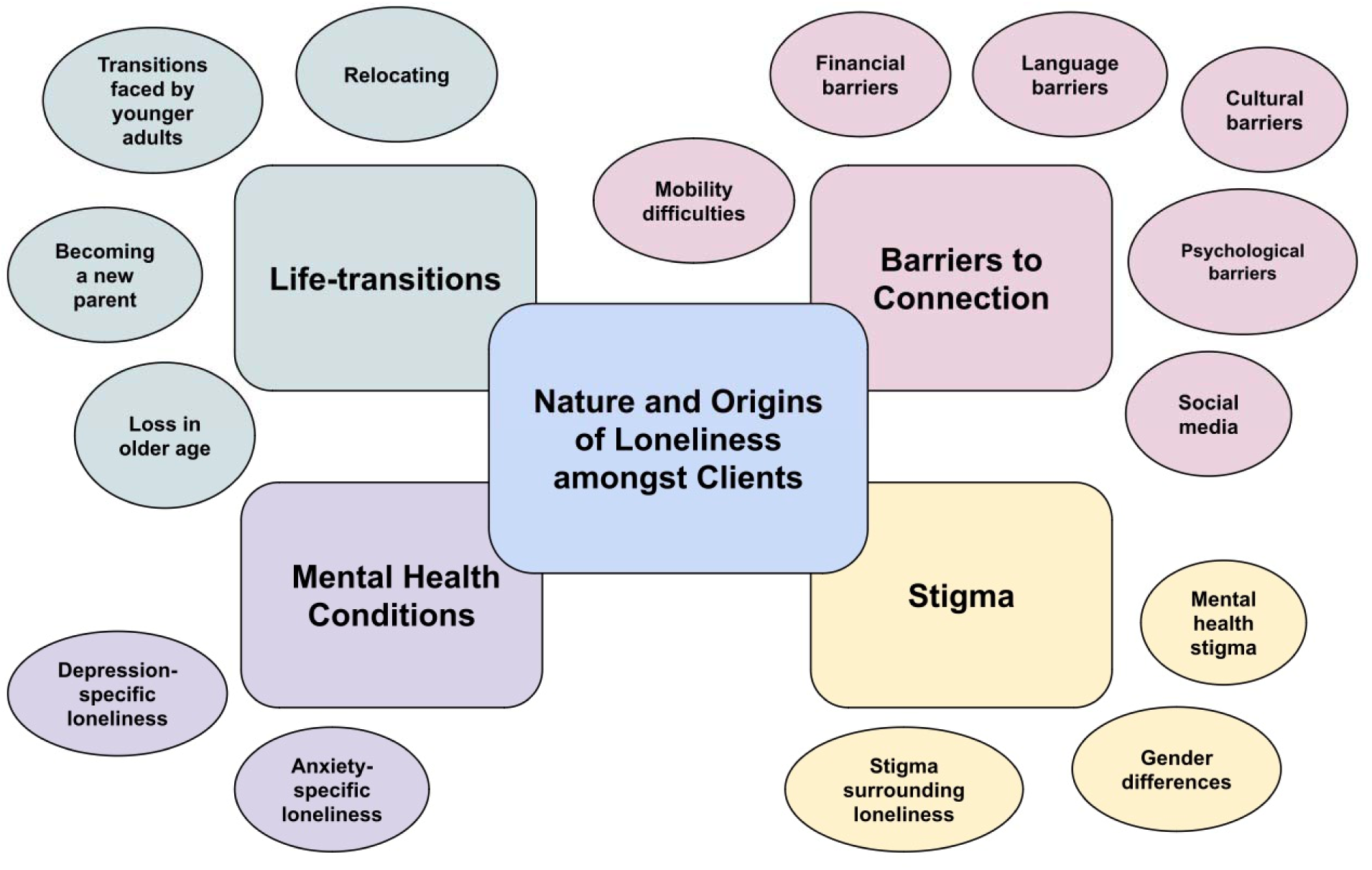
Diagram mapping key findings.

### Theme 1: Life-transitions

Therapists believed that many of their clients experience loneliness. Particularly, they reported that life transitions were linked to clients’ feelings of loneliness.

#### 1.1 Relocating

According to therapists, immigrating alone to the UK triggered feelings of loneliness amongst some clients. This was primarily due to the loss of social network, and difficulties in building new connections or communities:

> “*people that have immigrated to the UK… that sense of community, which they find to be supportive, and a big part of their lives, is lost.” [p1]*

> *“you don’t have much family because they have come alone… if they can find those communities then it is helpful. And some do not find it. So they miss home so much.” [p7]*

In particular, international students were brought up across many therapists to experience loneliness due to their relocation:

> *“students who have just started university… from maybe having quite a lot of friends and quite a big social circle where they grew up in their hometown and they’re moving to university. It can feel quite alienating, suddenly moving into a massive city where you don’t know anybody.” [p15]*

#### 1.2 Transitions faced by younger adults

Younger clients were also described to be facing unique transitions, from transitioning into university, to *(“finding [themselves] in society”)*. Particularly, these transitions were marked by the difficulties in finding *(“the right support systems”)* and peers that they could identify with:

> *“students, big transition, big changes, there’s a lot of loneliness at university or even at school if you don’t really identify with your peers… it’s a matter of finding your own people.” [p11]*

> *“by 18 you’re grown up, you come out, you’re in university, finding yourself in society… they have found themselves lonely… it’s not easy to find same wavelength of people.” [p7]*

#### 1.3 Becoming a parent

Some therapists noted how loneliness were experienced amongst new mothers, transitioning away from a *(“stable”)* social network due to their increased caregiving demands:

> *“primary caregiver, and often it is the mother, you’re no longer going to work and having that social network, that sort of stable contact, that connection that you’re used to having. And it can be quite isolating at home alone with your baby, so that’s another period of big change.” [p5]*

#### 1.4 Loss in older age

Older adults were also brought up by therapists to experience loneliness, due to the significant transitions experienced in their stage of lives. This includes *(“retirement”)* and the losing of partners and friends:

> *“When someone retires… they perhaps used to have a network, colleagues at work, and they no longer have that.” [p5]*

> *“suddenly, after having a fulfilled life, they are in their twilight years, and they’ve lost their partners.” [p7]*

### Theme 2: Mental health conditions

#### 2.1 Depression-specific loneliness

Many therapists pointed out loneliness to be a product of clients’ depression. Depressive symptoms of anhedonia, low mood and energy limited clients’ motivation to reach out to others or engage in social interactions, contributing to their feelings of loneliness and social isolation:

> *“I do see a lot of loneliness in especially people experiencing low mood… they start being quite withdrawn and isolated from people.” [p9]*

> *“not feeling like you have the energy or the motivation to go out and do some of these things [social interaction].” [p15]*

Feelings of loneliness associated with depression were also described by therapists as chronic in some clients, arising from cycles of low mood and withdrawal. Therapists noted that clients who persistently disengaged from social contact often lost touch with their social networks and became increasingly disconnected from others:

> *“it can be quite chronic, especially for people who are experiencing low mood, because when they are in that cycle of depression, they tend to withdraw from people, and as a result they don’t really meet up with their friends as often, they don’t really talk to people about what’s been going on, and then they feel quite disconnected from what other people are doing.” [p4]*

Apart from being a product of depression, loneliness was described by therapists as a factor that could cause or exacerbate depression symptoms amongst clients, given the fundamental human needs for social connection and belonging:

> *“It really correlates to clients with depression. I think a lot of the time the depression stems from loneliness.” [p8]*

> *“we have this real desire, don’t we, as humans to connect, and to feel supported, and to belong? And if that’s missing then of course it impacts on our mood and can lead us to feeling depressed.” [p5]*

#### 2.2 Anxiety-specific loneliness

Due to constant worries of others’ perception of oneself, clients with social anxiety were often mentioned by therapists to avoid socialising, feeding into their feelings of loneliness:

> *“clients who are experiencing social anxiety may sometimes say ‘I feel lonely’, because they do avoid going out, meeting people, socialising due to their anxiety of meeting people, worrying what other people are going to think of them, are they going to judge them.” [p9]*

Loneliness was also described to be linked with other types of anxiety. For instance, clients with generalised anxiety disorder might still socialise, although not able to be present and fully connect with others.

> *“with anxiety it’s like you still socialise… but you’re not necessarily getting the same enjoyment or interest out of it. Perhaps you’re so in your head, not really present because you’re worrying so much.” [p11]*

One therapist also mentioned that clients’ feelings of loneliness might be *(“further intensified”)* when others failed to understand their panic symptoms:

> *“feeling quite alone or lonely when they experienced let’s say a panic attack but their partner…doesn’t understand what that means.” [p4]*

Nonetheless, compared to depression, many therapists described loneliness as less associated with anxiety amongst clients. One therapists also mentioned that loneliness associated with anxiety might be more transient in nature:

> “*I don’t see a lot of how anxiety leads to loneliness… it’s not as common or prominent as clients experiencing low mood or depression.” [p9]*

> *“for people with anxiety or stress, it tends to be quite transient.” [p4]*

### Theme 3: Barriers to connection

Both practical and psychological barriers to connection were observed by therapists to contribute to clients’ feeling of loneliness.

#### 3.1 Mobility difficulties

Clients with physical health difficulties, particularly those affecting mobility, were often described to have experienced loneliness due to being housebound. This issue was prevalent amongst elderly clients:

> “*when someone loses their mobility, and they’re no longer even leaving the house…it leads to kind of really quite severe isolation.” [p5]*

> *“elderly population… people with mobility issues, not necessarily being able to leave your house and go out… to go see a friend or a family member.” [p15]*

#### 3.2 Financial barriers

From *(“go[ing] to the pub),* to joining *(“groups… hobbies… a gym”)*, or a *(“community group”)*, high cost of living in cities like London, together with financial difficulties faced by clients were believed to limit their ability to go out and socialise with people:

> *“Money, the cost-of-living crisis. People can’t afford to go to the pub, they can’t afford to go out and socialise.” [p8]*

> *“the asylum-seekers and the refugees… they have such limited money that they’re not going to spend… £4 on a return tube journey to get somewhere where they could go to a community group.” [p15]*

#### 3.3 Language barriers

A few therapists pointed out how difficulties speaking English in the UK might limit clients’ communications with others and their ability to *(“express”)* themselves or to form friendships:

> *“many patients that have difficultly speaking English… it makes it then difficult for them to understand certain humour, build certain relationships.” [p14]*

> *“language barrier, so they’re not able to reach out… you’re not able to talk and you’re not able to express.” [p7]*

One therapist also pointed out how language barriers might reduce accessibility to organisations made for minority groups, despite their existence in the community:

> *“there’s good organisations that try to reach out to minority groups… but it can definitely be an obstacle, especially if English isn’t your first language” [p11]*

#### 3.4 Cultural barriers

Many therapists described a sense of isolation experienced amongst clients who came from a non-English background. For instance, coming from (*“a collectivist culture”*), (*“Asian or ethnic minority backgrounds”*), some clients may *(“struggle to find themselves within the Western culture”*). The difference in *(“social cues, social constructs, social rules”)* also made it difficult for clients to *(“connect or communicate”)* with others, leading to feelings of loneliness:

> *“they feel like they’re an outcast or they feel quite isolated from the general community around them or the western culture itself.” [p9]*

> *“if you’re not a big drinker… culturally, we are quite keen on alcohol… certain lifestyle factors prevent people from being social.” [p14]*

In particular, one therapist highlighted that superficial interactions are characteristic of the British culture, as described by their clients. This hindered them in forming meaningful connections with others, which was *(“a contrast to experiences that people might have”)* where they *(“want that situation where they open up, and they share things”)*:

> *“They have described that British people, they don’t really go beyond superficial interactions unless they know you from childhood… people don’t go beyond that, and that makes them feel very isolated, very lonely.” [p1]*

London as a city was also described by some therapists as (*“lonely”*), (*“isolating”*) and (*“anonymous”*). (*“people come and go out of London quite quickly”)*, which might contrast the *(“very close-knit community”)* that their clients might be used to:

> *“London is very lonely. It’s a very highly compacted city but… When it comes to actual real connection it’s quite difficult.” [p14]*

#### 3.5 Psychological barriers

Even when practical barriers to connection were not present, many accounts of therapists suggest how clients might feel psychologically disconnected, (*“not able to communicate”*) or (*“can’t really be themselves”*):

> *“on the other hand, there’s people that actually have lots of people around them, and they don’t really engage with others, and they do feel lonely and isolated.” [p1]*

This might be due to clients’ upbringing or past negative experiences, or (“low self-esteem”):

> *“maybe someone has grown up learning that it’s bad or unhelpful or risky to rely on others and they naturally tend to keep themselves to themselves, don’t open up and don’t rely on anyone because perhaps they’ve had bad experiences in the past.” [p12]*

> *“the concept of low self-esteem… they’ll start to isolate themselves because they don’t want to be a burden… those preconceived ideas of not feeling worthy enough to receive that social support from other people.” [p15]*

#### 3.6 Social media

Some therapists mentioned that social media might contribute to loneliness, especially amongst younger clients. Despite being “connected” online, their interactions remain superficial, where clients *(“are not able to express”)*, or *(“share [their] sense of self”)*. Real-life connections might also drift away as a result:

> *“They are stuck within the imaginary world… that doesn’t help you to relate to people… slowly, slowly, people who they used to meet… have drifted away, and they’ve gotten themselves sort of locked up into this new… online friends… and you’re not able to express – these are strangers.”*

The constant comparison with others on social media might also exacerbate feelings of loneliness, mentioned by a few therapists:

> *“a factor of comparison… the quality of relationships isn’t really there… You just see people do things, but you don’t really interact, it’s more feelings of envy, of comparison, of inadequacy.” [p6]*

Nonetheless, one therapist mentioned that social media could also come with benefits, through the normalisation of loneliness and mental health:

> *“they might explore it [loneliness] a bit more and be more open about it just because… the internet… which normalises a lot of loneliness in society… the mental health content on social media, there’s a lot of therapists on social media.” [p11]*

### Theme 4: Stigma

#### 4.1 Mental health stigma

Many therapists highlighted the experience of mental health stigma amongst clients:

> *“It’s not the proper thing to do, to open up too much or share things.” [p1]*

> *“If you talk about it, it means you are weak.” [p13]*

Some therapists described how people around clients might *(“step away”)* from them due their mental health symptoms, or displayed *(“judgements”)* towards clients through their words or actions, which could intensify clients’ feelings of isolation and loneliness:

> *“the extreme reactions, the ups and downs. They don’t know what to do or maybe people have busy lives so they kind of think: I’m just going to step away from this person.” [p3]*

> “*even if they talk about it [real feelings], maybe someone didn’t help but by saying ‘you shouldn’t feel this’ and made them feel worse.” [p13]*

A few therapists described how clients might self-isolate, due to fears of being *(“judged”)* by or being a *(“burden”)* to others:

> *“fear of being ridiculed, fear of being excluded, fear of being judged, because of what they want to share, what’s happening in their lives, so in a way keeping up with pretences is a factor.” [p1]*

> *“They stop talking to people because they’re scared that it’s going to be a burden on other people.” [p15]*

In particular, one therapist pointed out that older age might play a role in this internalised stigma, which could intensify their existing sense of loneliness:

> *“if they share that with people, then they are afraid that they might be seen as pathetic or not an adult, or ‘look at this person, they’re a certain age and they can’t really feel good’” [p1]*

#### 4.2 Gender differences

Many therapists explicitly highlighted the societal expectations around *(“man needing to act strong”).* Despite having *(“a good social network”),* some male clients’ were described to struggle with sharing their feelings or opening up about their mental health problems, interacting at *(“surface level relationships”):*

> *“there’s a lot of stigma around the man needing to act strong and not be very vocal with what they’re talking and what they’re sharing. And they usually are quite closed off.” [p14]*

> *“I’m seeing a client at the moment who’s male and he has lots of friends, he socialises a lot, but he doesn’t talk about anything of meaning or his emotions or the fact that he’s having problems with his mental health. So he feels really lonely and isolated because they’re not having these conversations because he feels so ashamed to.” [p8]*

Contrarily, female clients were perceived by some therapists as *(“less emotionally alone”)* due to having more support systems that allowed them for self-expression:

> *“Whereas women… They tend to report more frequently having a small circle of close friends who know what’s going on in their life and they talk to about their mental health.” [p8]*

### 4.3 Stigma surrounding loneliness

Other than stigma surrounding mental health symptoms, therapists described the very feeling of loneliness to be stigmatised in our society, something their clients felt shameful about:

> *“I think there’s a lot of stigma about being alone” [p11]*

> *“I think loneliness can be quite a touchy conversation. It can be something that people are really deeply embarrassed about, and feel abnormal in.” [p14]*

Particularly, feelings of shame seemed to arise in their clients’ when they compared their (lack of) social lives with others:

> *“if somebody is feeling lonely… they’re gonna be over-thinking the loneliness-‘Why am I different?’ ‘Why don’t I have this social network.?’ ‘Why is everybody else happy?’” [p14]*

For clients who had a social circle, guilt might be experienced where they blamed themselves for feeling lonely:

> *“shame and guilt…‘Everyone is OK, why I’m feeling this?’ ‘Oh I have a family and friend why I’m feeling lonely? Is something wrong with me?’” [p13]*

> *“they actually do have a social network and they feel guilty about still feeling lonely.” [p14]*

Therapists also mentioned that stigma around loneliness was present during therapy sessions, where clients found it hard to *(“admit”)* their feelings of loneliness:

> *“They don’t describe themselves feeling lonely… they find it hard to admit that, even to me in the sessions.” [p1]*

> *“There might even be an element of shame to saying ‘I’m lonely… it can be difficult to come forward from the client themselves.” [p10]*

## Discussion

To our knowledge, this study is the first to examine loneliness amongst NHS TTad clients with depression and anxiety from therapists’ perspectives. Findings align with existing literature on loneliness, showing that loneliness in NHS TTad clients arose from both external (e.g., life transitions, financial difficulties) and internal factors (e.g., low self-esteem), and can persist despite social connections due to structural, cultural, and psychological barriers.

Therapists perceived loneliness as varied across age, gender, and disorder type, where stigma around mental health and loneliness may perpetuate clients’ feelings of loneliness.

According to therapists’ accounts, various life transitions might trigger loneliness amongst clients. This aligns with previous research identifying events like relocating (44), starting university (45), retiring (46), widowhood (47), and becoming a parent (48) as triggers for loneliness. Following the Social Identity Model of Identity Change (49), life transitions might trigger feelings of loneliness amongst clients, due to a disruption in social groups and loss of social identity. In particular, age-normative events experienced by both younger and older clients were notable triggers of loneliness, which echoes with research that shows a curvilinear relationship between loneliness and age (50), and how age-related life events are linked with changes in social networks (51).

Moreover, therapists also highlighted the close and cyclical link between loneliness and depressive symptoms (e.g., anhedonia, social withdrawal). This echoes with two studies that found chronic loneliness to increase the risk of major depression (52,53), a recent network analysis which shows anhedonia to be a key link between depression and loneliness (54), and two longitudinal studies that display a bidirectional relationship between loneliness and depression (55,56).

Contrarily, anxiety disorders’ relationship with loneliness appeared less prominent from our therapists’ accounts. This aligns with a recent systematic review of longitudinal studies, which found mixed results regarding the association between loneliness and anxiety (57).

Nonetheless, in our study, when clients’ anxiety-related loneliness was probed, social anxiety and its associated fear and avoidance of social interaction were frequently noted by the therapists.

This aligns with Lim et al.’s (29) quantitative finding that social anxiety predicts higher loneliness over time. Still, further research is needed to explore the relationship between different types of anxiety disorders (e.g., phobic, generalized anxiety) and loneliness, as each may have unique associations.

As captured in theme 3, therapists highlighted external barriers to social connection, such as mobility restrictions and financial difficulties, which limited clients’ ability to maintain relationships. This aligns with research showing higher loneliness amongst those with physical disabilities and financial challenges (58–60). Our study further adds qualitative insight into how such barriers impact clients’ social engagement. Importantly, theme 3 revealed that clients’ loneliness extended beyond physical social connections. Despite having people around them, therapists noted how cultural, language, and psychological barriers might still hinder clients’ ability to connect with others. In particular, clients not originally from the UK were perceived to be prone to loneliness by therapists, due to cultural barriers, which echoes with other qualitative research capturing difficulties of international students integrating into Western cultures (61,62). What our study further pinpoints is that language barrier might limit clients’ ability to express themselves, directly feeding into their feelings of loneliness. Refugees and asylum-seekers are also a group highlighted by many therapists to experience loneliness, due to financial, cultural, and language barriers, as well as loss of social networks from relocation. This aligns with Deighton-O’Hara’s qualitative findings (63), which identified similar challenges amongst refugees in the UK, such as social isolation, cultural barriers, and difficulties with identity and services. Given the unique challenges faced by this group, including uncertain legal status and limited access to resources, further research on their loneliness, especially within clinical populations, is warranted.

Therapists also described how macro-level factors, such as stigma and societal expectations, contributed to clients’ loneliness. Our findings align with existing research distinguishing between public and self-stigma. Public stigma, involving societal endorsement of stereotypes and exclusionary behaviour toward individuals with mental illness (64), was reflected in therapists’ accounts of clients feeling distanced or shamed by others. Self-stigma, the internalisation of these attitudes leading to reduced self-esteem and social withdrawal (65), was similarly evident in clients who feared burdening or being judged by others. This echoes previous qualitative findings that young people with depression experienced both forms of stigma, resulting in withdrawal (65). The current study extends this understanding by suggesting that similar processes may also operate amongst individuals with anxiety disorders.

Furthermore, therapists noted that male clients appeared particularly vulnerable to loneliness associated with mental health stigma. This observation aligns with theories of hegemonic masculinity, which emphasise cultural ideals of toughness and emotional restraint in men (66). Such expectations may underlie therapists’ reports of male clients concealing mental health struggles and maintaining only superficial interactions. These accounts echo findings from qualitative research with UK-based men showing reluctance to disclose loneliness and difficulty forming close connections (67). Although meta-analytic evidence suggests minimal gender differences in loneliness overall (68), our findings highlight gendered mechanisms through which men may experience and express loneliness, warranting further investigation.

Lastly, therapists described that loneliness itself could be stigmatised amongst clients. In the subtheme *“Stigma surrounding loneliness,”* therapists observed clients expressing self-stigma—feeling ashamed or guilty about their loneliness and concealing it, even within therapy. This might stem from perceived public stigma, or fear of discrimination for being lonely (69).

These accounts align with UK-based studies reporting that fear of others’ reactions and self-blame create barriers to expressing loneliness (70,71). However, therapists’ accounts rarely included concrete instances of public stigma, consistent with studies that found limited or mixed evidence of actual public stigma of lonely individuals, despite the presence of self-stigma (71–74). Further research is needed to clarify how perceived and actual stigma interact in shaping lonely individuals’ experiences.

## Strengths and limitations

The current study addressed a key gap in the literature by providing an in-depth exploration of loneliness amongst NHS TTad clients from the perspective of a key stakeholder group (therapists). This focus is particularly important given that NHS TTad serves a large population with depression and anxiety—two of the most prevalent mental health conditions in England. Particularly, as NHS TTad therapists manage diverse caseloads varying in demographic and clinical characteristics, their accounts captured a wide range of loneliness experiences and associated patterns (e.g., demographic differences). The sample’s diversity further strengthened the study, as participating therapists varied in age, service location, job role (with equal numbers of high- and low-intensity therapists), and level of experience. Methodological rigour was enhanced through multiple reflexive strategies, having interviews conducted by two researchers, regular team discussions, diary keeping, and independent coding by a second researcher.

Despite its contributions, this study has several limitations. All recruited therapists were based in London, which may limit the transferability of findings to other English regions, given the city’s distinctive characteristics such as high living costs and greater ethnic diversity. Our sample also consisted predominantly of female and White therapists, which, although reflects the demographics of the NHS TTad workforce (staff are predominantly female and/or White) (75), could still have restricted gender- or culture-specific insights—e.g., potential differences in interpretations of loneliness or in therapeutic dynamics. Finally, although reflexivity was embedded throughout the analytic process, neither NHS TTad therapists nor clients were directly involved in data analysis, which might have constrained the depth of interpretive insight.

## Research and clinical implications

Future research should directly involve interviewing NHS TTad service users to corroborate and expand upon the present findings. Including both therapists and service users in data analysis could further enhance the depth and relevance of interpretations. Recruiting participants from diverse regions across England would also allow examination of potential geographical variations in loneliness experiences. Additionally, several themes identified in this study warrant deeper investigation. Future work should explore how different types of anxiety disorders relate to loneliness, and whether cyclical patterns similar to those observed in depression exist. It would also be valuable to explore whether clients’ loneliness experiences are influenced by the severity of depression and anxiety symptoms. Examining loneliness amongst specific demographic groups, such as asylum seekers is also important.

Exploring the relationship between social media use and loneliness, especially amongst younger people is crucial, given the mixed results in current literature (76) and the nuanced patterns observed in our findings. Particularly, research should examine which aspects of social media use contribute to loneliness and which promote connectedness. Gender differences also merit closer examination, considering the unique challenges faced by each gender, e.g., females’ potential loneliness related to child-rearing and males’ difficulties in acknowledging or reporting loneliness. In addition, the underexplored area of public stigma surrounding loneliness should be addressed through both qualitative and quantitative approaches.

Therapists’ accounts highlighted loneliness as common amongst NHS TTad clients, cutting across diverse demographic and diagnostic characteristics. Clinically, this underscores the importance of therapists recognising that some clients may experience loneliness alongside, or as part of, their mental health difficulties. Assessing and addressing loneliness may therefore be valuable in both case formulation and treatment. Future research should examine how loneliness can be effectively identified and managed within, or outside of the NHS TTad services.

Finally, tackling stigma around both mental health and loneliness remain essential, as stigma continues to hinder help-seeking. Broader interventions at societal, educational, and policy levels—including targeted anti-stigma campaigns—are needed to foster more supportive environments for individuals experiencing loneliness or mental health difficulties.

## CRediT Author Statement

**Cheuk Hei Peony Chung**: Conceptualisation, Formal Analysis, Investigation, Visualisation, Writing – Original Draft Preparation

**Brynmor Lloyd-Evans**: Conceptualisation, Project Administration, Funding Acquisition, Supervision, Validation, Writing – Review & Editing

**Theodora Stefanidou**: Conceptualisation, Data Curation, Funding Acquisition, Investigation, Methodology, Project Administration, Resources, Supervision, Validation, Writing – Review & Editing

**Sampath Rajapakse**: Validation

## Data Availability

All data produced in the present study are available upon reasonable request to the authors

## References

1. World Health Organisation. World Health Organisation. Mental disorders [Internet]. 2023 [cited 2025 Sep 27]. Available from: https://www.who.int/news-room/fact-sheets/detail/mental-disorders.

2. Institute for Health Metrics and Evaluation. Disease, injury, and risk factsheets [Internet]. 2024 [cited 2025 Sep 27]. Available from: https://www.healthdata.org/research-analysis/diseases-injuries-risks/factsheets.

3. World Health Organisation. Disability-adjusted life years (DALYs) [Internet]. 2024 [cited 2025 Sep 27]. Available from: https://www.who.int/data/gho/indicator-metadata-registry/imr-details/158.

4. McDaid D, Park AL, Davidson G, John A, Knifton L, McDaid S, et al. The economic case for investing in the prevention of mental health conditions in the UK [Internet]. 2022 [cited 2025 Sep 27]. Available from: https://pure.qub.ac.uk/en/publications/the-economic-case-for-investing-in-the-prevention-of-mental-healt.

5. McManus S, Bebbington PE, Jenkins R, Brugha T. Mental health and wellbeing in England: the adult psychiatric morbidity survey 2014. Leeds: NHS Digital; 2016.

6. Clark DM. Implementing NICE guidelines for the psychological treatment of depression and anxiety disorders: the IAPT experience. Int Rev Psychiatry. 2011;23(4):318–27.

7. NHS England. NHS Talking Therapies, for anxiety and depression, Workforce [Internet]. 2024 [cited 2025 Sep 27]. Available from: https://www.england.nhs.uk/mental-health/adults/nhs-talking-therapies/workforce/.

8. NHS Digital. NHS Talking Therapies, for anxiety and depression, Annual reports, 2024–25 [Internet]. 2024 [cited 2025 Sep 27]. Available from: https://digital.nhs.uk/data-and-information/publications/statistical/nhs-talking-therapies-for-anxiety-and-depression-annual-reports/2024-25.

9. O’Donnell J, Pybis J, Bacon J. Counselling in the third sector: to what extent are older adults accessing these services and how complete are the data third sector services collect measuring client psychological distress? Counselling and Psychotherapy Research. 2021 Feb 26;21(2).

10. Bhavsar V, Jannesari S, McGuire P, MacCabe JH, Das-Munshi J, Bhugra D, et al. The association of migration and ethnicity with use of the Improving Access to Psychological Treatment (IAPT) programme: a general population cohort study. Social Psychiatry and Psychiatric Epidemiology. 2021 Feb 16;56(11):1943–56.

11. Green SA, Honeybourne E, Chalkley SR, Poots AJ, Woodcock T, Price G, et al. A retrospective observational analysis to identify patient and treatment-related predictors of outcomes in a community mental health programme. BMJ Open. 2015 May 1;5(5):e006103.

12. Verbist IL, Allsopp K, Huey D, Varese F. Frequency and impact of childhood sexual and physical abuse on people using IAPT services. Br J Clin Psychol. 2021 Nov;60(4):504–12.

13. Clark DM, Canvin L, Green J, Layard R, Pilling S, Janecka M. Transparency about the outcomes of mental health services (IAPT approach): an analysis of public data. The Lancet. 2018 Feb;391(10121):679–86. .

14. Goddard E, Wingrove J, Moran P. The impact of comorbid personality difficulties on response to IAPT treatment for depression and anxiety. Behaviour Research and Therapy. 2015 Oct 1;73:1–7.

15. Thomlinson R, Muncer S, Dent H. Comorbidity between PTSD and anxiety and depression: implications for IAPT services. Archives of Depression and Anxiety. 2017 May 30;3(1):14–7.

16. Peplau LA, Perlman D. Perspectives on loneliness. Loneliness: A sourcebook of current theory, research and therapy. 1982 Jul 7:1–8.

17. Cacioppo S, Grippo AJ, London S, Goossens L, Cacioppo JT. Loneliness: clinical import and interventions. Perspect Psychol Sci. 2015;10(2):238–49.

18. Hawkley LC, Browne MW, Cacioppo JT. How can I connect with thee? Let me count the ways. Psychol Sci. 2005;16(10):798–804.

19. Hawkley LC, Hughes ME, Waite LJ, Masi CM, Thisted RA, Cacioppo JT. From social structural factors to perceptions of relationship quality and loneliness: the Chicago health, aging, and social relations study. J Gerontol B Psychol Sci Soc Sci. 2008;63(6):S375–84.

20. Chau AKC, So SH, Sun X, Zhu C, Chiu CD, Chan RCK, et al. The co-occurrence of multidimensional loneliness with depression, social anxiety and paranoia in non-clinical young adults: a latent profile analysis. Front Psychiatry. 2022;13:931558.

21. Diehl K, Jansen C, Ishchanova K, Hilger-Kolb J. Loneliness at universities: determinants of emotional and social loneliness among students. Int J Environ Res Public Health. 2018;15(9):1865.

22. Wolters NE, Mobach L, Wuthrich VM, Vonk P, Van der Heijde CM, Wiers RW, et al. Emotional and social loneliness and their unique links with social isolation, depression and anxiety. J Affect Disord. 2023;329:207–17.

23. Raut NB, Singh S, Subramanyam AA, Pinto C, Kamath RM, Shanker S. Study of loneliness, depression and coping mechanisms in elderly. J Geriatr Ment Health. 2014;1(1):20–7.

24. Meltzer H, Bebbington P, Dennis MS, Jenkins R, McManus S, Brugha TS. Feelings of loneliness among adults with mental disorder. Soc Psychiatry Psychiatr Epidemiol. 2013 Jan;48(1):5–13.

25. Erzen E, Çikrikci Ö. The effect of loneliness on depression: a meta-analysis. Int J Soc Psychiatry. 2018;64(5):427–35.

26. Park C, Majeed A, Gill H, Tamura J, Ho RC, Mansur RB, et al. The effect of loneliness on distinct health outcomes: a comprehensive review and meta-analysis. Psychiatry Res. 2020;294:113514.

27. Wang J, Mann F, Lloyd-Evans B, Ma R, Johnson S. Associations between loneliness and perceived social support and outcomes of mental health problems: a systematic review. BMC Psychiatry. 2018;18:156.

28. Dour HJ, Wiley JF, Roy-Byrne P, Stein MB, Sullivan G, Sherbourne CD, et al. Perceived social support mediates anxiety and depressive symptom changes following primary care intervention. Depress Anxiety. 2014;31(5):436–42.

29. Lim MH, Rodebaugh TL, Zyphur MJ, Gleeson JF. Loneliness over time: the crucial role of social anxiety. J Abnorm Psychol. 2016;125(5):620–30.

30. Maes M, Nelemans SA, Danneel S, Fernández-Castilla B, Van den Noortgate W, Goossens L, et al. Loneliness and social anxiety across childhood and adolescence: multilevel meta-analyses of cross-sectional and longitudinal associations. Dev Psychol. 2019;55(7):1548–65.

31. Lasgaard M, Goossens L, Elklit A. Loneliness, depressive symptomatology, and suicide ideation in adolescence: cross-sectional and longitudinal analyses. J Abnorm Child Psychol. 2011;39:137–50.

32. McHugh Power J, Hannigan C, Hyland P, Brennan S, Kee F, Lawlor BA. Depressive symptoms predict increased social and emotional loneliness in older adults. Aging Ment Health. 2020;24(1):110–8.

33. Gao S, Assink M, Cipriani A, Lin K. Associations between rejection sensitivity and mental health outcomes: a meta-analytic review. Clin Psychol Rev. 2017;57:59–74.

34. Allen NB, Badcock PB. The social risk hypothesis of depressed mood: evolutionary, psychosocial, and neurobiological perspectives. Psychol Bull. 2003;129(6):887–913.

35. Cacioppo JT, Hawkley LC, Thisted RA. Perceived social isolation makes me sad: 5-year cross-lagged analyses of loneliness and depressive symptomatology in the Chicago Health, Aging, and Social Relations Study. Psychol Aging. 2010;25(2):453–63.

36. Birken M, Chipp B, Shah P, Olive RR, Nyikavaranda P, Hardy J, et al. Exploring the experiences of loneliness in adults with mental health problems: a participatory qualitative interview study. PLoS One. 2023;18(3):e0280946.

37. Ludwig KA, Brandrett B, Lim MH, Mihas P, Penn DL. Lived experience of loneliness in psychosis: a qualitative approach. J Ment Health. 2022;31(4):543–50.

38. Ikhtabi S, Pitman A, Toh G, Birken M, Pearce E, Johnson S. The experience of loneliness among people with a “personality disorder” diagnosis or traits: a qualitative meta-synthesis. BMC Psychiatry. 2022;22(1):130.

39. Achterbergh L, Pitman A, Birken M, Pearce E, Sno H, Johnson S. The experience of loneliness among young people with depression: a qualitative meta-synthesis of the literature. BMC Psychiatry. 2020;20:1–23.

40. Achterbergh L, Pitman A, Birken M, Pearce E, Sno H, Johnson S. The experience of loneliness among young people with depression: a qualitative meta-synthesis of the literature. BMC Psychiatry. 2020;20:1–23.

41. Hennink M, Kaiser BN. Sample sizes for saturation in qualitative research: a systematic review of empirical tests. Soc Sci Med. 2022;292:114523.

42. Braun V, Clarke V. Conceptual and design thinking for thematic analysis. Qual Psychol. 2022;9(1):3–26.

43. Wilson C, Janes G, Williams J. Identity, positionality and reflexivity: relevance and application to research paramedics. Br Paramed J. 2022;7(2):43–9.

44. Evans O, Cruwys T, Cárdenas D, Wu B, Cognian AV. Social identities mediate the relationship between isolation, life transitions, and loneliness. Behav Change. 2022;39(3):191–204.

45. Cruwys T, Ng NW, Haslam SA, Haslam C. Identity continuity protects academic performance, retention, and life satisfaction among international students. Appl Psychol. 2021;70(3):931–54.

46. Haslam C, Steffens NK, Branscombe NR, Haslam SA, Cruwys T, Lam BCP, et al. The importance of social groups for retirement adjustment: evidence, application, and policy implications of the social identity model of identity change. Soc Issues Policy Rev. 2019;13(1):93–124.

47. Sheftel MG, Margolis R, Verdery AM. Life events and loneliness transitions among middle-aged and older adults around the world. J Gerontol B Psychol Sci Soc Sci. 2024;79(1):gbad149.

48. Seymour-Smith M, Cruwys T, Haslam SA. More to lose? Longitudinal evidence that women whose social support declines following childbirth are at increased risk of depression. Aust N Z J Public Health. 2021;45(4):338–43.

49. Haslam C, Haslam SA, Jetten J, Cruwys T, Steffens NK. Life change, social identity, and health. Annu Rev Psychol. 2021;72(1):635–61.

50. Mund M, Freuding MM, Möbius K, Horn N, Neyer FJ. The stability and change of loneliness across the life span: a meta-analysis of longitudinal studies. Pers Soc Psychol Rev. 2020;24(1):24–52.

51. Wrzus C, Hänel M, Wagner J, Neyer FJ. Social network changes and life events across the life span: a meta-analysis. Psychol Bull. 2013;139(1):53–80.

52. Martín-María N, Caballero FF, Lara E, Domènech-Abella J, Haro JM, Olaya B, et al. Effects of transient and chronic loneliness on major depression in older adults: a longitudinal study. Int J Geriatr Psychiatry. 2021;36(1):76–85.

53. Cacioppo JT, Hawkley LC, Ernst JM, Burleson M, Berntson GG, Nouriani B, et al. Loneliness within a nomological net: an evolutionary perspective. J Res Pers. 2006;40(6):1054–88.

54. Yang M, Wei W, Ren L, Pu Z, Zhang Y, Li Y, et al. How loneliness linked to anxiety and depression: a network analysis based on Chinese university students. BMC Public Health. 2023;23(1):2499.

55. Choi EY, Yoon H, Lee HY, Cho J. Bidirectional relationships between loneliness and depressive symptoms among older adults: a 12-year population-based prospective study. Int Psychogeriatr. 2020;32(4):457–67.

56. Ren L, Mo B, Liu J, Li D. A cross-lagged regression analysis of loneliness and depression: a two-year trace. Eur J Dev Psychol. 2022;19(2):198–212.

57. Stefanidou T, Evlat G, Gray H, Lagan K, Harju-Seppänen J, Osullivan M, et al. Associations between loneliness and outcomes of common mental disorders (CMDs): a systematic review of longitudinal studies. medRxiv [Preprint]. 2025 Aug.

58. Rokach A, Lechcier-Kimel R, Safarov A. Loneliness of people with physical disabilities. Soc Behav Pers. 2006;34(6):681–700.

59. LV=. A third of people struggling financially also feel lonely [Internet]. 2022 [cited 2025 Sep 27]. Available from: https://www.lvadviser.com/knowledge-centre/news-hub/a-third-of-people-struggling-financially-also-feel-lonely.

60. Kung CS, Pudney SE, Shields MA. Economic gradients in loneliness, social isolation and social support: evidence from the UK Biobank. Soc Sci Med. 2022;306:115122.

61. Newsome LK, Cooper P. International students’ cultural and social experiences in a British university: “Such a hard life [it] is here”. J Int Students. 2016;6(1):195–215.

62. O’Dea X, Stern J. Cross-cultural integration through the lens of loneliness: a study of Chinese direct entry students in the United Kingdom. Qual Res Educ. 2022;11(3):203–29.

63. Deighton-O’Hara L. Refugee and migrant well-being in the UK: post-migration challenges for those in exile. A review of the literature. MIDIRS Midwifery Dig. 2018;28(2):249–54.

64. Corrigan PW, Watson AC. Understanding the impact of stigma on people with mental illness. World Psychiatry. 2002;1(1):16–20.

65. Ben-Zeev D, Young MA, Corrigan PW. DSM-V and the stigma of mental illness. J Ment Health. 2010;19(4):318–27.

66. Connell RW. Masculinities. Berkeley: University of California Press; 1995.

67. Ratcliffe J, Kanaan M, Galdas P. Reconceptualising men’s loneliness: an interpretivist interview study of UK-based men. Soc Sci Med. 2023;332:116129.

68. Maes M, Qualter P, Vanhalst J, Van den Noortgate W, Goossens L. Gender differences in loneliness across the lifespan: a meta-analysis. Eur J Pers. 2019;33(6):642–54.

69. Barreto M, van Breen J, Victor C, Hammond C, Eccles A, Richins MT, et al. Exploring the nature and variation of the stigma associated with loneliness. J Soc Pers Relatsh. 2022;39(9):2658–79.

70. Co-op Foundation. All our emotions are important: breaking the silence about youth loneliness. Manchester: Co-op Foundation; 2018.

71. Sawdon K, et al. Loneliness stigma rapid evidence assessment (REA) [Internet]. London: UK Government; 2023 [cited 2025 Sep 27]. Available from: https://www.gov.uk/government/publications/research-exploring-the-stigma-associated-with-loneliness/loneliness-stigma-rapid-evidence-assessment-rea.

72. Christensen PN, Kashy DA. Perceptions of and by lonely people in initial social interaction. Pers Soc Psychol Bull. 1998;24(3):322–9.

73. Kerr NA, Stanley TB. Revisiting the social stigma of loneliness. Pers Individ Dif. 2021;171:110482.

74. Lau S, Gruen GE. The social stigma of loneliness: effect of target person’s and perceiver’s sex. Pers Soc Psychol Bull. 1992;18(2):182–9.

75. NHS Benchmarking Network. Psychological Professions: Workforce Census in England at 31 March 2024. NHS England; 2025.

76. Zhang L, Li C, Zhou T, Li Q, Gu C. Social networking site use and loneliness: a meta-analysis. J Psychol. 2022;156(7):492–511.

