## Appendices for "The Nature and Origins of Loneliness amongst NHS Talking Therapies Clients— A Qualitative Study from Therapists’ Perspective"

**Appendix A** - **Consolidated criteria for reporting qualitative studies (COREQ): 32-item checklist**

Developed from: Tong A, Sainsbury P, Craig J. Consolidated criteria for reporting qualitative research (COREQ): a 32-item checklist for interviews and focus groups. International journal for quality in health care. 2007 Dec 1;19(6):349-57.

| Item No | | Guide Questions/Description | Reported on Page # |  |  |
| --- | --- | --- | --- | --- | --- |
| Domain 1: Research team and reflexivity | | | |  |  |
| Personal Characteristics | | | |  |  |
| 1. Interviewer/ facilitator | | Which author/s conducted the interview or focus group? | 9 |  |  |
| 2. Credentials | | What were the researcher’s credentials? E.g., PhD, MD | 10 |  |  |
| 3. Occupation | | What was their occupation at the time of the study? | 10 |  |  |
| 4. Gender | | Was the researcher male or female? | 10 |  |  |
| 5. Experience and training | | What experience or training did the researcher have? | 10 |  |  |
| Relationship with participants | | | |  |  |
| 6. Relationship established | | Was a relationship established prior to study commencement? | N/A |  |  |
| 7. Participant knowledge of the interviewer | | What did the participants know about the researcher? e.g. personal goals, reasons for doing the research? | N/A |  |  |
| 8. Interviewer characteristics | | What characteristics were reported about the interviewer/facilitator? e.g. Bias, assumptions, reasons and interests in the research topic | 10 |  |  |
|  | Domain 2: study design | | |  |  |
|  | Theoretical framework | | |  |  |
|  | 9. Methodological orientation and Theory | What methodological orientation was stated to underpin the study? e.g. grounded theory, discourse analysis, ethnography, phenomenology, content analysis | 9 |  |  |
|  | Participant selection | | |  |  |
|  | 10. Sampling | How were participants selected? e.g., purposive, convenience, consecutive, snowball | 8 |  |  |
|  | 11. Method of approach | How were participants approached? e.g., face-to-face, telephone, mail, email | 8 |  |  |
|  | 12. Sample size | How many participants were in the study? | 11 |  |  |
|  | 13. Non-participation Setting | How many people refused to participate or dropped out? Reasons? | 8 |  |  |
|  | 14. Setting of data collection | Where was the data collected? e.g., home, clinic, workplace | 9 |  |  |
|  | 15. Presence of nonparticipants | Was anyone else present besides the participants and researchers? | N/A |  |  |
|  | 16. Description of sample | What are the important characteristics of the sample? e.g. demographic data, date | 11 |  |  |
|  | Data collection | | |  | No |
|  | 17. Interview guide | Were questions, prompts, and guides provided by the authors? Was it pilot tested? | 9, Appendix B |  |  |
|  | 18. Repeat interviews | Were repeat interviews carried out? If yes, how many? | N/A |  |  |
|  | 19. Audio/visual recording | Did the research use audio or visual recording to collect the data? | 9 |  |  |
|  | 20. Field notes | Were field notes made during and/or after the interview or focus group? | N/A |  |  |
|  | 21. Duration | What was the duration of the interviews or focus group? | 9 |  |  |
|  | 22. Data saturation | Was data saturation discussed? | 8 |  |  |
|  | 23. Transcripts returned | Were transcripts returned to participants for comment and/or correction? | N/A |  |  |
|  | Domain 3: analysis and findings | | |  |  |
|  | Data analysis | | |  |  |
|  | 24. Number of data coders | How many data coders coded the data? | 9 |  |  |
|  | 25. Description of the coding tree | Did the authors provide a description of the coding tree? | N/A |  |  |
|  | 26. Derivation of themes | Were themes identified in advance or derived from the data? | 9 |  |  |
|  | 27. Software | What software, if applicable, was used to manage the data? | 9 |  |  |
|  | 28. Participant checking | Did participants provide feedback on the findings? | N/A |  |  |
|  | Reporting | | |  |  |
|  | 29. Quotations presented | Were participant quotations presented to illustrate the themes/findings? Was each quotation identified? e.g., participant number | 13-21 |  |  |
|  | 30. Data and findings consistent | Was there consistency between the data presented and the findings? | 12-21 |  |  |
|  | 31. Clarity of major themes | Were major themes clearly presented in the findings? | 12-21 |  |  |
|  | 32. Clarity of minor themes | Is there a description of diverse cases or a discussion of minor themes? | 12-25 |  |  |

### Appendix B – Topic guide for NHS TTad staff

**A. Information about you**

- Gender (Male; Female; Prefer not to say; I describe my gender with another term - please state:………)
- Age (18-30; 31-50; 51+; prefer not to say)
- Ethnicity (White/White British; Black/Black British; Asian/Asian British; Mixed ethnic groups; prefer not to say; I describe my gender with another term – please state:….)
- NHS TTad organisation you work for (which part of the country)
- How long have you worked in NHS TTad services? (Less than 6 months – 2 years; 2-5 years; 6-10 years; more than 10 years)
- How long have you worked in mental health services generally?
- Job title:

*Preamble*

***This is a flexible guide to help build a conversation. Not all questions will need to be asked and may not be relevant to all interviewees. Interviewers will paraphrase question to best suit the person they are speaking with.*** ***The purpose of this guide is to explore people’s views and experiences of working with NHS TTad clients that experience loneliness, the extent of the problem, what’s already in place and what else can be done to support lonely clients as well as the mechanisms through which loneliness might affect treatment and engagement as well as treatment outcomes.***

  Start recording

**A. Extent of loneliness for NHS TTad clients**

1. Is loneliness an issue for many of your clients?

Prompts:

- Do they talk about feeling lonely or isolated?
- How do you know that they feel lonely? (Emerges naturally; therapists asking explicitly)
- Do these feelings tend to be chronic or transient or a mixture? Do you notice any differences among clients who are recently lonely compared to those experiencing chronic issues?

2. Is loneliness and social connections something you often explore with clients?

Prompts:

- Is exploring loneliness or social connections part of your job? Do you have adequate time to do it?
- What’s the best way to identify NHS TTad clients who are lonely?
- Any standard questions you ask to find out about loneliness?
- Would it be helpful to use loneliness screening tools as part of the routine data collection in NHS TTad?

3. From your experience working with clients, what are the biggest factors that people experience loneliness?

Prompts:

- Social isolation; Mental health problems; Relationship factors; Social factors; Personality related factors; health problems; post-covid pandemic (adolescents; work from home; psychological impact) relationship with self, historical causes, geographic relocation, economic/financial factors
- To what extent do the people you see tend to experience loneliness due to mental health or things going on their life?

4. From your experience, are any client groups more prone to loneliness?

Prompts:

- Demographics: age, marital status, new mothers, same sex attracted people, education, employment, ethnic communities, language barriers, disabilities, cost of living and socioeconomic issues e.g., transport costs and infrastructure, alcohol culture? Non-drinkers? Cultural differences?
- Certain psychological disorders? I.e., Depression, social anxiety, substance misuse, agoraphobia, other phobias, OCD, paranoia

**B. Staff response to loneliness**

5. Can you offer any specific support to people that are lonely or lacking social connections? Is it part of your job to address loneliness?

Prompts:

- If anything, are you able to offer people any kind of support for their feelings of loneliness?
- How do you talk about it with your clients? (Language and attitude)
- How do you feel talking about loneliness?

6. Has anything in your training and experience helped you in addressing loneliness? Do you feel you would benefit from more training?

Prompts

- Do you have resources to signpost people to help with social connections? Or does your service do that? Links with social prescribing

7. Is there anything else you would like to do as part of your NHS TTad role to support clients expressing a high degree of loneliness?

8. Is NHS TTad the right setting to support lonely clients with depression and anxiety? Is current NHS TTad help enough to support clients who have depression and anxiety and are lonely? (within the service)

Prompts:

- What are the barriers?
- If you think it’s not the right setting what can help?
- The service enhanced with more psychological or social connections provision?

9. What other types of support would be helpful to support lonely clients? (And sequence of how to do it) (other services)

Prompts:

- Referrals to other services
- Social interventions? Social prescribing – connectors. How would that fit into NHS TTad?
- Psychological interventions?
- CBT module for loneliness?
- Community Interventions?

**C. Loneliness mechanisms**

10. What are the mechanisms by which loneliness might affect treatment engagement for lonely clients?

Prompts

- Appointment attendance, therapeutic alliance

11. What are the mechanisms by which loneliness might affect treatment outcomes for lonely clients? (Depression and anxiety outcomes)

12. Use Cacioppo’s framework to introduce different types of loneliness before asking the below questions.

*Loneliness is a complex construct that includes three related facets or dimensions: 1) Intimate loneliness; 2) Relational loneliness; and 3) Collective loneliness (Hawkley et al., 2005; Hawkley, Gu, Luo, & Cacioppo, 2012).*

*“Intimate loneliness refers to the perceived absence of a significant someone (e.g., a spouse), that is, a person one can rely on for emotional support during crises, who provides mutual assistance, and who affirms one’s value as a person.”*

*“Relational loneliness refers to the perceived presence/absence of quality friendships or family connections.”*

*“Collective loneliness refers to a person’s valued social identities or “active network” (e.g., group, school, team, or national identity) wherein an individual can connect to similar others at a distance in the collective space.”*

a. From your experience, which type of loneliness is more prevalent among NHS TTad clients?
b. Do certain types of loneliness affect more certain groups of clients?
c. Do we need different interventions for different types of loneliness?

13. Is there anything else you would like to tell us about how loneliness affects NHS TTad treatment engagement and outcomes?

** We will write a report summarising the findings of the study. Ask participants at the end of the interview if they would like to receive a copy of the report. The findings will also be submitted for publication in a peer-reviewed journal. Participants will not be personally identifiable in any of these reports from the study.**
